# Clinical evidence of variable proton relative biological effectiveness in locally advanced non–small-cell lung cancer patients

**DOI:** 10.1101/2025.09.10.25335510

**Authors:** Tien T. Tang, Yulun He, Kevin McCoy, Antony Adair, Pablo Yepes, Carol C. Wu, Carlos E. Cardenas, Peter Balter, Julianne Pollard-Larkin, David B. Flint, Christine B. Peterson, Zhongxing Liao, Radhe Mohan, Kristy Brock

## Abstract

**Background and Purpose:** To derive and validate the proton variable relative biological effectiveness (RBE) from CT imaging changes in patients with locally advanced non–small-cell lung cancer.

**Materials and Methods:** We retrospectively analyzed data for patients previously treated on a prospective randomized trial with standard fractionated intensity-modulated photon therapy (IMPT) or passive scattering proton therapy (PSPT). CT image density change (IDC) was calculated in the normal ipsilateral lung between the planning and follow-up CT. Using IDC as the clinical evidence of response, we correlated dose–IDC relationships by fitting a modified Lyman–Kutcher–Burman NTCP model for each treatment modality. RBE was calculated with corresponding planned dose for the IMRT patients and planned physical dose for PSPT patients from the fitted NTCP model. To validate the measured clinical radiographic-based RBE values, we benchmarked the measured RBE values with those calculated from the McNamara, Wedenberg, Mairani, and Flint empirical models. In addition, we also fitted the McNamara and Wedenberg models using the measured RBE, LET_d_, and IDC values.

**Results:** The IDC-based RBE values ranged from 1.3-2.3. Linear regression comparing the radiographic-based RBE value with the empirical models, returned R^2^ values of 0.025-0.72. The fitted McNamara and Wedenberg models returned pseudo-R^2^ values of 0.95 and 0.85, respectively.

**Conclusion:** RBE values can be computed from CT image density changes as the clinical endpoint and validated using published empirical models. This is the first study to demonstrate the potential of using a clinical radiographic-based endpoint to improve our understanding of the biological effects of proton therapy.

## Introduction

For patients with locally advanced non–small-cell lung cancer (LA-NSCLC), standard of care is concurrent chemoradiation therapy which can often lead to radiation-induced toxicity such as radiation pneumonitis (1, 2). The conventional radiation modality is intensity-modulated radiation therapy (IMRT). However, IMRT can deliver a considerable amount of radiation to surrounding healthy tissues due to exit dose.

An alternative is proton therapy, which offers superior normal-tissue sparing and reduced toxicity risk (3). These advantages are due to the Bragg peak phenomenon, in which protons, upon entering the patient, slow down continuously at an increasing rate and deposit most of their energy at the end of their range, thus causing the most biological damage at the end of their range and spare tissues beyond the distal edge of the beam (4, 5). Therefore, proton therapy can deposit a considerably smaller dose in normal tissues outside the target compared to photon therapy. However, studies indicate that these two treatment modalities have similar efficacy and toxicity rates. A randomized phase II trial comparing passive scattering proton therapy (PSPT) and IMRT for LA-NSCLC found that although PSPT had better dose conformality—reducing the lung volume receiving at least 5 or 10 Gy—it did not significantly lower rates of local failure or grade 3 pneumonitis (1). This suggests that the expected tissue-sparing benefits of proton therapy have not translated into improved clinical outcomes, implying a gap in our understanding of the underlying biological effects of proton-induced damage in the lung.

The relative biological effectiveness (RBE) is the ratio of the dose of a reference radiation source, usually photons, needed to achieve a biological endpoint versus the dose of an ion radiation source needed to achieve the same endpoint. A constant proton RBE of 1.1 remains the consensus in clinical practice however clinical trials have not confirmed the anticipated superior tissue sparing of proton therapy (1, 6–8). A possible factor that may contribute to this disparity is a fixed-RBE–weighted dose in lung was maintained for PSPT and increased RBE that was not accounted for during treatment planning could have contributed to the increased toxicity.

The hypothesis that RBE can vary has prompted the development of many phenomenological RBE models using empirical data from *in vitro* cell studies (9–12). In general, these models assume a linear-quadratic mechanism of cell killing and incorporate parameters such as photon dose, proton dose (physical), linear energy transfer (LET), and alpha/beta ratio (α/β), as well as unique fitting parameters that are strongly dependent on dose and LET. However, these models do not adequately reflect the normal tissue response in patients and lack *in vivo* verification. RBE values derived directly from patient data could better reveal the clinically relevant biological effects of protons. Many studies have demonstrated the feasibility of determining normal tissue response with localized image-based dose–response models (13–17). However, there is currently no established *in vivo* RBE model using clinical endpoints.

In this study, we used the image density change (IDC) between the planning computed tomography (CT) and the post-treatment CT as a measurable clinical endpoint for RBE, defined as the ratio of photon and proton doses causing the same level of IDC. We validated the image-derived RBE values against those obtained using established empirical RBE models. The objective of this study is to leverage clinical image changes to determine the variable dose-RBE relationship that will offer further insight into proton-induced biological damage in patients.

## Methods

### Patients

Under an Institutional Review Board–approved protocol, images from 106 patients with LA-NSCLC who previously received standard fractionation of 2 Gy, IMRT (n = 60) or PSPT (n = 46) to 60–74 Gy (RBE = 1.1 for PSPT) in a randomized trial were used in this study, Table 1 (1). All participants proved written informed consent. We recomputed the planned dose for IMRT patients (D_x_) in a commercial treatment planning system (RayStation 11B DTK, RaySearch Laboratories, Stockholm, Sweden). For PSPT patients, we recomputed the planned physical dose (D_p_) by dividing the biological dose by 1.1 and the dose-averaged LET (LET_d_) using a clinically commissioned track-repeating Monte Carlo algorithm (18). Dose and LET_d_ characteristics for the IMRT and PSPT cohorts are shown in Supplementary Figure 1 and 2. For each patient, a contrast-free, free-breathing 4-dimensional CT was acquired for treatment planning. In addition, a contrast-free, free-breathing follow-up CT was acquired approximately 4 months post RT (IMRT: 4.1 [range: 1.3–8] months vs. PSPT: 3.6 [range: 0.4–7.3] months; *P* = 0.14, unpaired Student t-test).

**Table 1.**
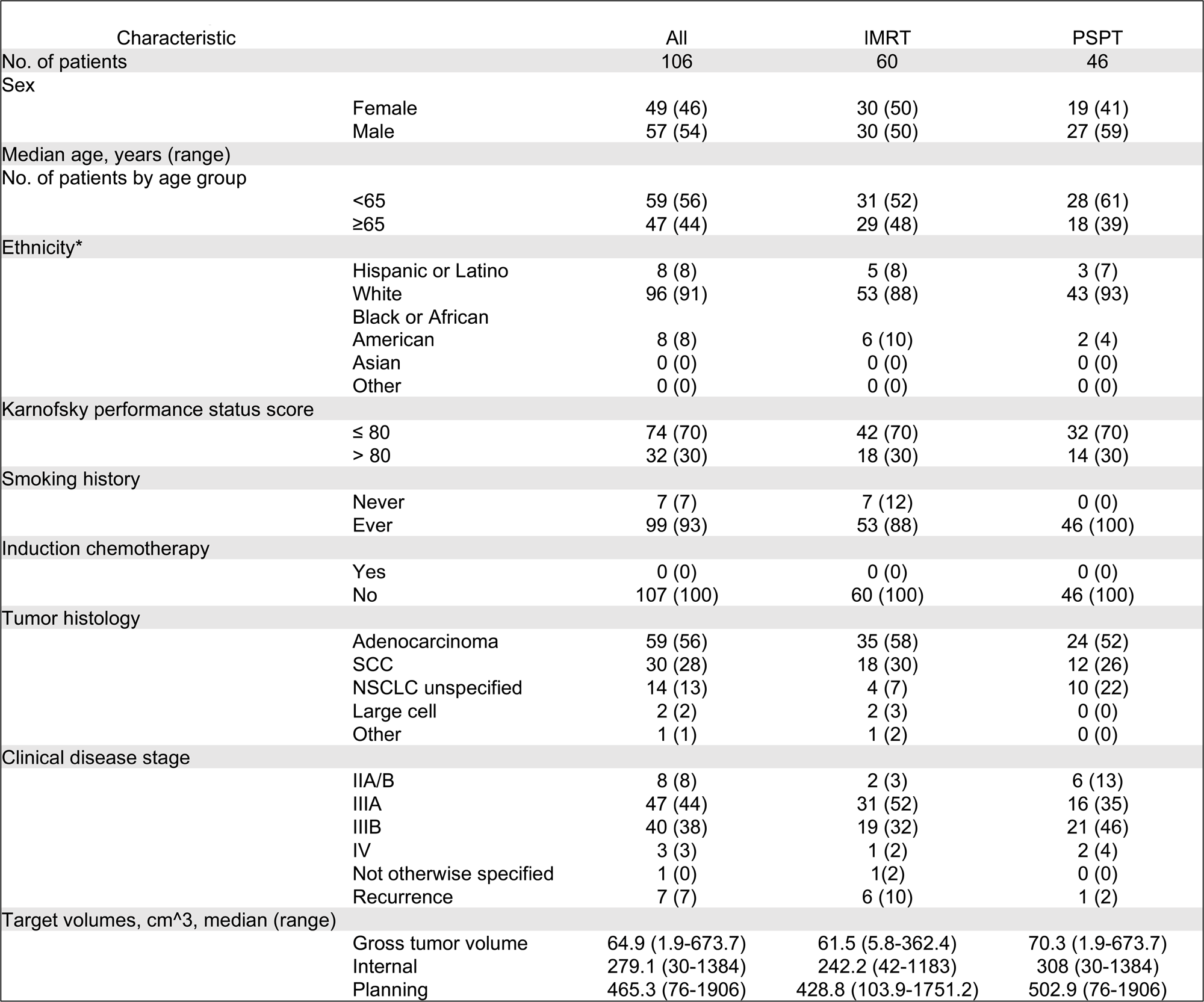
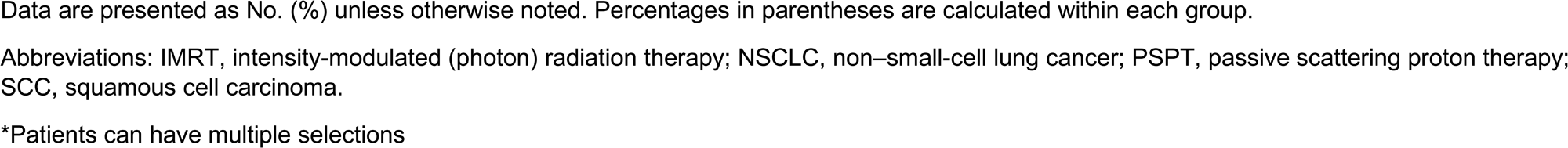
Patient Demographics.

| **Table 1. Patient Demographics** |  |  |  |  |
| --- | --- | --- | --- | --- |
| Characteristic |  | All | IMRT | PSPT |
| No. of patients |  | 106 | 60 | 46 |
| Sex |  |  |  |  |
|  | Female | 49 (46) | 30 (50) | 19 (41) |
|  | Male | 57 (54) | 30 (50) | 27 (59) |
| Median age, years (range) |  |  |  |  |
| No. of patients by age group |  |  |  |  |
|  | <65 | 59 (56) | 31 (52) | 28 (61) |
|  | ≥65 | 47 (44) | 29 (48) | 18 (39) |
| Ethnicity* |  |  |  |  |
|  | Hispanic or Latino | 8 (8) | 5 (8) | 3 (7) |
|  | White | 96 (91) | 53 (88) | 43 (93) |
|  | Black or African American | 8 (8) | 6 (10) | 2 (4) |
|  | Asian | 0 (0) | 0 (0) | 0 (0) |
|  | Other | 0 (0) | 0 (0) | 0 (0) |
| Karnofsky performance status score |  |  |  |  |
|  | ≤ 80 | 74 (70) | 42 (70) | 32 (70) |
|  | > 80 | 32 (30) | 18 (30) | 14 (30) |
| Smoking history |  |  |  |  |
|  | Never | 7 (7) | 7 (12) | 0 (0) |
|  | Ever | 99 (93) | 53 (88) | 46 (100) |
| Induction chemotherapy |  |  |  |  |
|  | Yes | 0 (0) | 0 (0) | 0 (0) |
|  | No | 107 (100) | 60 (100) | 46 (100) |
| Tumor histology |  |  |  |  |
|  | Adenocarcinoma | 59 (56) | 35 (58) | 24 (52) |
|  | SCC | 30 (28) | 18 (30) | 12 (26) |
|  | NSCLC unspecified | 14 (13) | 4 (7) | 10 (22) |
|  | Large cell | 2 (2) | 2 (3) | 0 (0) |
|  | Other | 1 (1) | 1 (2) | 0 (0) |
| Clinical disease stage |  |  |  |  |
|  | IIA/B | 8 (8) | 2 (3) | 6 (13) |
|  | IIIA | 47 (44) | 31 (52) | 16 (35) |
|  | IIIB | 40 (38) | 19 (32) | 21 (46) |
|  | IV | 3 (3) | 1 (2) | 2 (4) |
|  | Not otherwise specified | 1 (0) | 1 (2) | 0 (0) |
|  | Recurrence | 7 (7) | 6 (10) | 1 (2) |
| Target volumes, cm<sup>3</sup>, median (range) |  |  |  |  |
|  | Gross tumor volume | 64.9 (1.9-673.7) | 61.5 (5.8-362.4) | 70.3 (1.9-673.7) |
|  | Internal | 279.1 (30-1384) | 242.2 (42-1183) | 308 (30-1384) |
|  | Planning | 465.3 (76-1906) | 428.8 (103.9-1751.2) | 502.9 (76-1906) |

Data are presented as No. (%) unless otherwise noted. Percentages in parentheses are calculated within each group.
Abbreviations: IMRT, intensity-modulated (photon) radiation therapy; NSCLC, non–small-cell lung cancer; PSPT, passive scattering proton therapy; SCC, squamous cell carcinoma.
\*Patients can have multiple selections

### Establishing the dose–response relationship

A commercially available biomechanical model–based deformable image registration (DIR) algorithm was used to deform the follow-up CT to the planning CT to compute IDC for each voxel in the normal, ipsilateral lung (19). Voxels were at least 2 mm from the boundaries of the lung and the contoured primary gross target volume (GTV) on the planning CT. All contours were reviewed by a board-certified radiologist (CW) and radiation oncologist (ZL).

To establish the dose–IDC correspondence for each voxel, we resampled D_x_, D_p_, and LET_d_ maps into 1.98 x 1.98 x 5.0 mm^3^ “super” voxels with a diagonal dimension of 5.7 mm to account for DIR uncertainty as recommended by AAPM TG-132 (20). For each patient, the mean (physical) dose and mean IDC were calculated for each dose bin (5-Gy intervals). For each bin, dose and IDC values were averaged within patient then averaged over patients. The averaged values were used to correlate voxel-level dose–IDC relationships by fitting the Lyman–Kutcher–Burman (LKB) normal-tissue complication probability (NTCP) model (21, 22). Please refer to supplementary material for additional information.

### Computing RBE values and fitting RBE models

The measured IDC-based RBE values were calculated from the fitted NTCP models by using the ratio of D_x_ to D_p_. To validate the measured RBE values, we compared them to RBE values derived from four empirical models: McNamara *et al.* (*9*), Wedenberg *et al.* (*23*), Mairani *et al.* (*24*), and Flint *et al.* (*25*). We also fitted the McNamara (9) and the Wedenberg (23) models with our dataset.

### Statistics

Statistical analyses were conducted using Prism 10 (GraphPad Software, Inc.). An unpaired Student t-test was used for statistical analyss between groups. To fit the NTCP model we used R (nls, R 4.4.3). A linear regression model was used to evaluate the relationship between the measured RBE values and those derived from the empirical models. To fit the McNamara and Wedenberg models we used python (curve_fit, v3.11.9). Model fit was assessed using the pseudo R^2^, which allows relative comparison of models fit to the same data set, and residual sum of squares values. Linear regression model was fitted to

## Results

In general, the distributions of dose and IDC between the radiation cohorts were similar, with no significant differences in the mean, standard deviation, or median value across patients (Supplementary Fig. 1). The only parameters that were significantly different between the cohorts were minimum dose (IMRT: 0.8±0.8Gy, PSPT: 0.102±0.08Gy, p<0.0001) and maximum dose (IMRT: 81±5Gy, PSPT: 77±6Gy, p=0.0004). This is to be expected as one of the key advantages of proton therapy is sparing of normal tissue and conformal irradiation of the tumor. An example of the dose and LET distribution is shown in Fig. 1.

**Figure 1.**
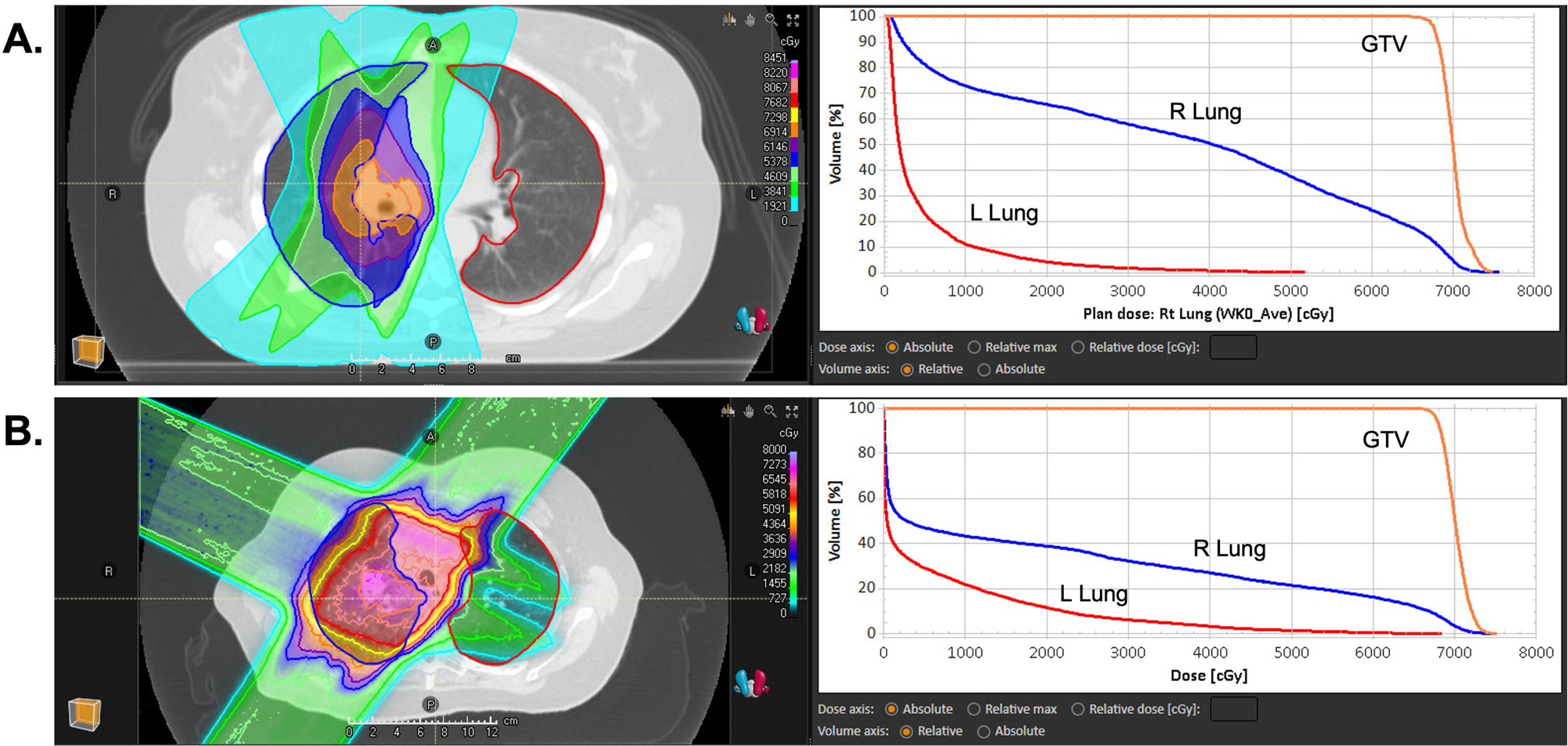
Treatment plan and corresponding dose volume histogram for the right lung, left lung and gross tumor volume (GTV) of an intensity-modulated photon therapy plan (A) and passive scattering proton therapy plan (B).

The LKB NTCP fitting results of IDC dose-response at the population level for IMRT patients and PSPT patients are shown in Fig. 2. PSPT had a higher estimated IDC than IMRT for a given dose bin except for the two highest dose bins. TD50, the dose for uniform irradiation which results in a 50% probability of max IDC, was higher for PSPT compared to IMRT. In general, the LKP NTCP model was a good fit for the dose–IDC relationship for both the IMRT (residual standard error (RSE) = 0.04, m: SE = 0.029, TD50: SE = 0.81) and PSPT (RSE = 0.08 m: SE = 0.083, TD50: SE = 1.49) cohort, suggesting that this model can be used for RBE modeling of clinical data at the population level. However, at the patient level, we found large patient variability for both cohorts in the IDC-dose response (Supplementary Fig. 3). To identify the lowest and highest IDC levels from the population RBE model, we compared the fitted curves. Because the PSPT curve was higher than the IMRT curve, we set the lowest IDC level at 20 HU (based on the PSPT curve) and set the highest IDC level at 115 HU (based on the IMRT curve). The D_x_, D_p_ and the estimated LET_d_ for each IDC level is given in Table 2. To demonstrate that RBE calculated based on IDC (RBE_IDC_) depends on the physical beam parameters similarly to RBE calculated based on cell survival, we graphed RBE as a function of LET_d_ and dose and found a very strong correlation between RBE_IDC_ and LET_d_ / D_p_ (p<0.0001) (Fig. 3A). This is consistent with the known dependencies of proton RBE with LET and dose, namely that RBE increases with increasing LET and decreases with increasing dose.

**Figure 2.**
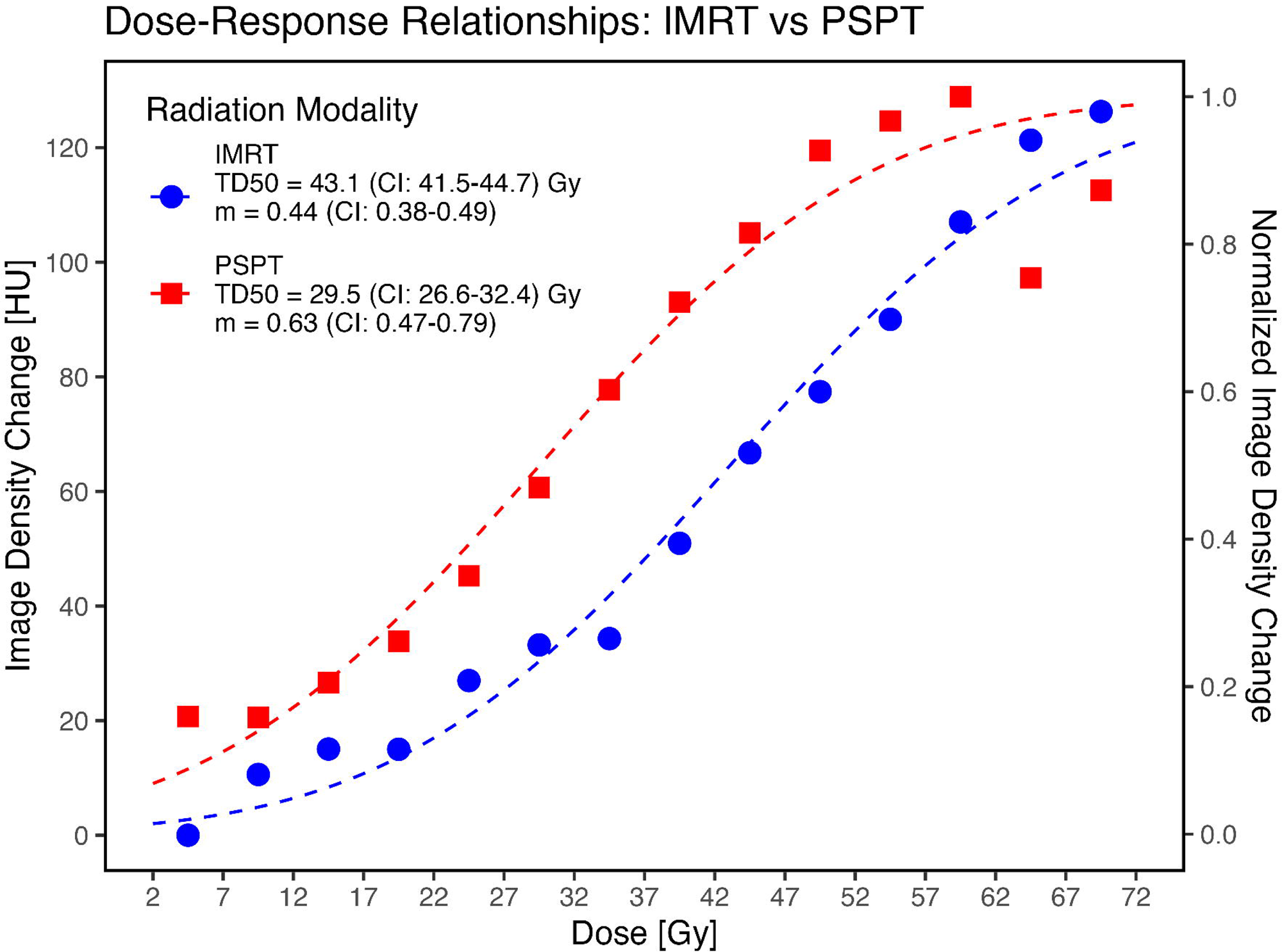
Dose–response modeling results for patients who received either passive scattering proton therapy (red square) or intensity-modulated photon therapy (blue circle). The mean image density change (IDC), mean dose, and IDC normalized for voxels in each dose bin are shown. The dashed lines represent the fitted models.

**Figure 3.**
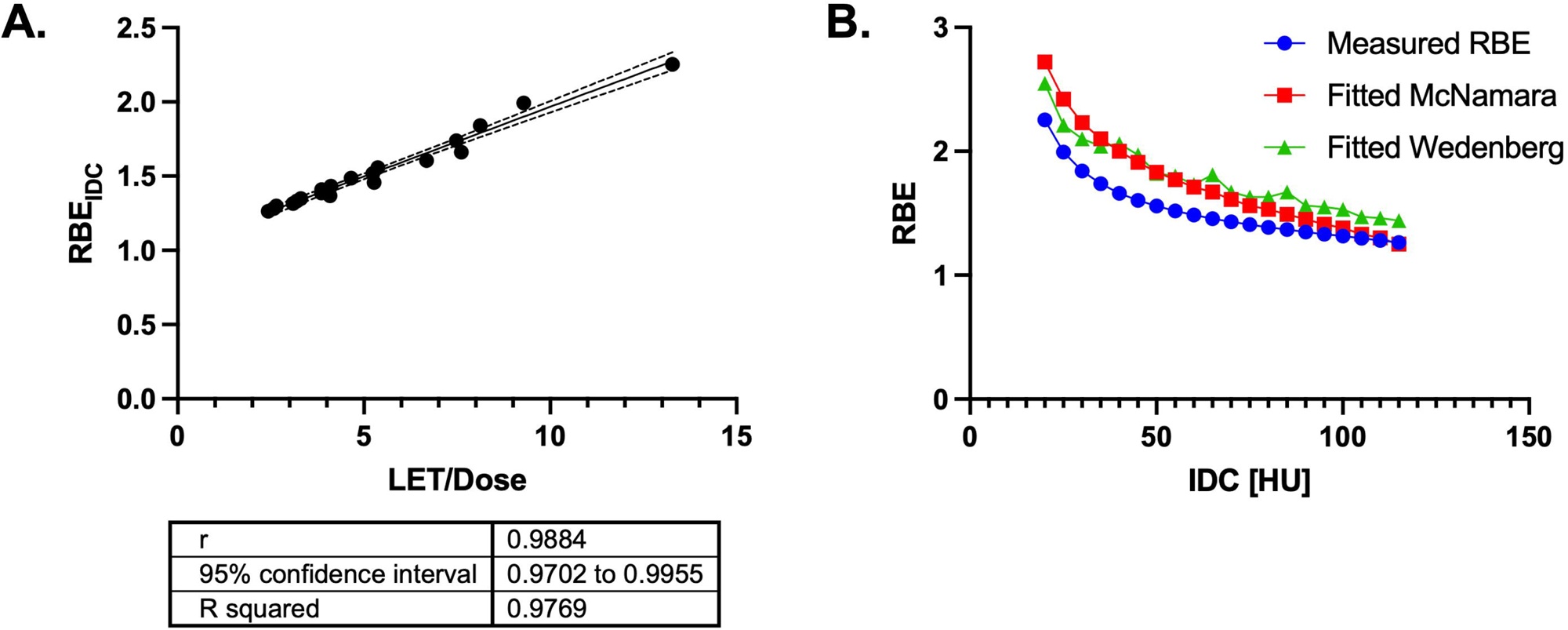
Relationship between the measured RBE and the ratio of LET and proton dose per fraction. The solid line is the best-fit linear regression line, and the dashed lines indicate the 95% confidence interval (A). Comparison of the measured RBE value to the fitted RBE values from the McNamara and Wedenberg model at different IDC levels (B).

**Table 2.** Results of the relative biological effectiveness (RBE) modeling to corresponding IDC levels.

| IDC [HU] | D<sub>x</sub> [Gy] | D<sub>p</sub> [Gy] | LET<sub>d</sub> [keV/μm] | Measured RBE (based on imaged response) | RBE fitted using McNamara model | RBE fitted using Wedenberg model |
| --- | --- | --- | --- | --- | --- | --- |
| 20 | 23.96 | 10.63 | 3.82 | 2.25 | 2.72 | 2.55 |
| 25 | 26.81 | 13.45 | 3.38 | 1.99 | 2.42 | 2.21 |
| 30 | 29.33 | 15.94 | 3.50 | 1.84 | 2.23 | 2.10 |
| 35 | 31.62 | 18.20 | 3.68 | 1.74 | 2.10 | 2.04 |
| 40 | 33.75 | 20.30 | 4.18 | 1.66 | 2.00 | 2.06 |
| 45 | 35.77 | 22.29 | 4.03 | 1.60 | 1.91 | 1.97 |
| 50 | 37.70 | 24.21 | 3.51 | 1.56 | 1.83 | 1.82 |
| 55 | 39.58 | 26.06 | 3.69 | 1.52 | 1.77 | 1.80 |
| 60 | 41.43 | 27.88 | 3.51 | 1.49 | 1.71 | 1.73 |
| 65 | 43.25 | 29.69 | 4.23 | 1.46 | 1.67 | 1.81 |
| 70 | 45.08 | 31.50 | 3.50 | 1.43 | 1.61 | 1.67 |
| 75 | 46.93 | 33.32 | 3.48 | 1.41 | 1.56 | 1.63 |
| 80 | 48.82 | 35.19 | 3.66 | 1.39 | 1.53 | 1.63 |
| 85 | 50.77 | 37.11 | 4.10 | 1.37 | 1.49 | 1.67 |
| 90 | 52.80 | 39.12 | 3.49 | 1.35 | 1.45 | 1.56 |
| 95 | 54.96 | 41.25 | 3.56 | 1.33 | 1.41 | 1.55 |
| 100 | 57.28 | 43.55 | 3.65 | 1.32 | 1.38 | 1.53 |
| 105 | 59.85 | 46.09 | 3.29 | 1.30 | 1.33 | 1.47 |
| 110 | 62.78 | 48.98 | 3.41 | 1.28 | 1.30 | 1.46 |
| 115 | 66.29 | 52.45 | 3.44 | 1.26 | 1.25 | 1.44 |

Abbreviations: IDC, image density change; D<sub>x</sub>, photon dose; D<sub>p</sub>, proton physical dose; LET, linear energy transfer.

We obtained the following estimated parameter values from fitting the McNamara model: p_0_ = 26.71 (SE = 68.41, 95% CI: −118.30,171.73), p_1_ = 1.76 (SE = 54.74, 95% CI: −114.28-117.81), p_2_ = 0.01 (SE = 739.20, 95% CI: −1567.03-1567.04), and p_3_ = −0.0011 (SE = 177.88, 95% CI: −377.09-377.09), with a pseudo-R^2^ of 0.95 and residual sum of squares of 0.35. When fitting the Wedenberg model, which is similar to the McNamara model but with only one free parameter, we obtained a parameter estimate of q = 16.57 (standard error = 1.34, 95% CI:13.76-19.38), with a pseudo-R^2^ of 0.85 and residual sum of squares of 1.17. Of the two models, the McNamara had a better fit (Figure 3B), but with higher uncertainty in the fitted free parameters. Comparison of the RBE values from the fitted McNamara and Wedenberg models showed the measured RBE values tend to predict lower RBE values for lower IDC values (Table 2).

To validate the measured radiographic-based RBE values, we compared them to the empirically derived RBE values calculated from four different models. Linear regression analysis demonstrated a positive correlation between the measured and empirical RBE values (R^2^ = 0.68, 0.72, 0.72, and 0.025 for the McNamara, Wedenberg, Flint, and Mairani models, respectively). We found that the measured RBE values showed a significant correlation with the empirical RBE values, except for the Mairani mode (Fig. 4). The measured RBE values were generally higher than the empirical RBE values. Specifically, at lower doses, the measured RBE values were found to be higher than those predicted by any of the four models (Supplementary Fig. 4).

**Figure 4.**
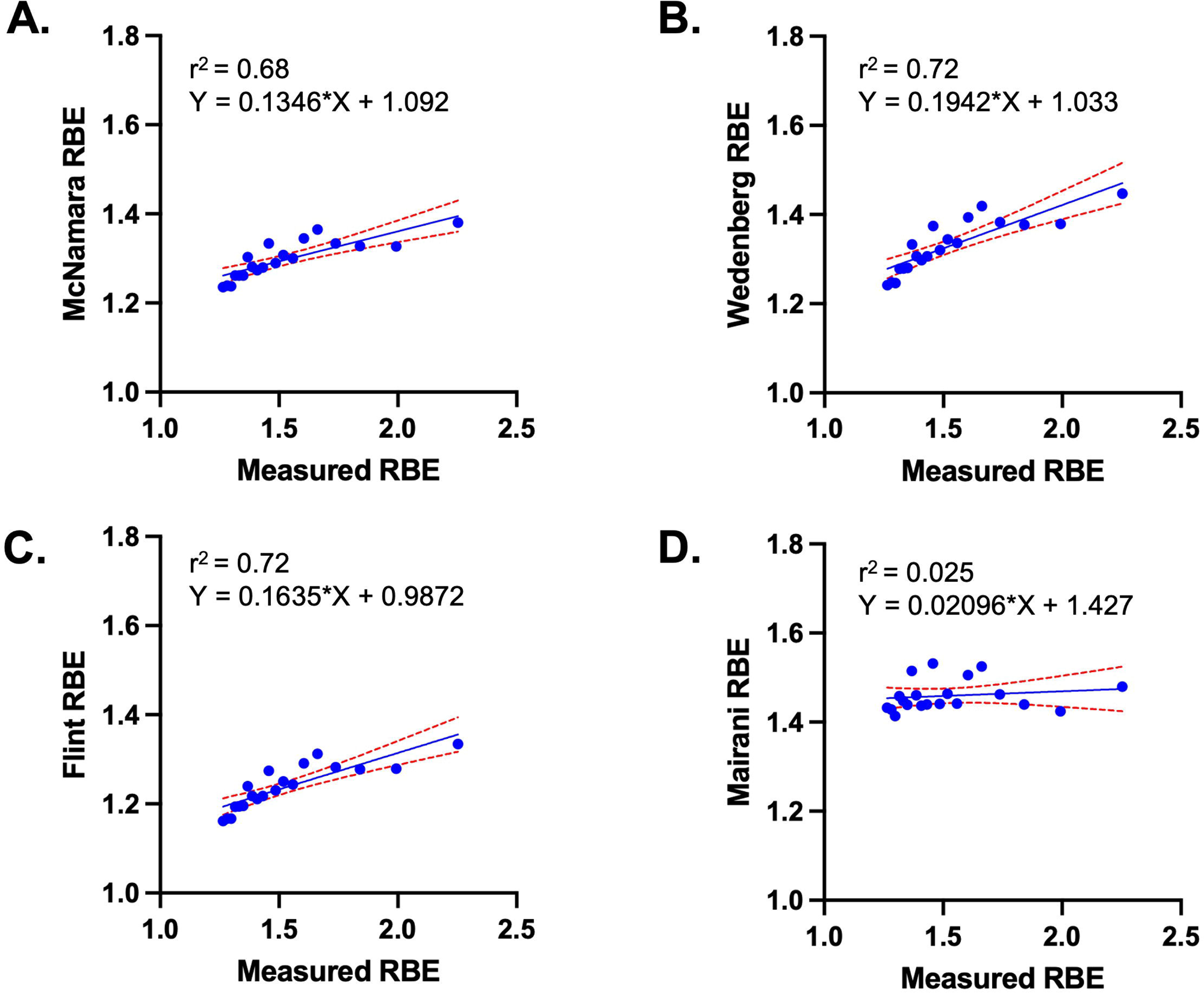
Comparison of the measured RBE with those calculated using the McNamara (A), Wedenberg (B), Mairani (C), and Flint model (D). The solid lines are the best-fit linear regression lines, and the dashed lines indicate the 95% confidence interval.

## Discussion

In this study, we introduced a novel approach to modeling proton RBE in which clinical imaging response in patients was used as the biological endpoint and demonstrated a variable dose–RBE relationship. The measured RBE values were also found to be comparable to values derived from four published empirical RBE models and have the same dependency with LET and dose as RBE calculated from *in vitro* studies. This is the first step towards the implementation of *in vivo* RBE modeling, and towards identifying noninvasive RBE endpoints.

Others have proposed the feasibility of using intensity changes on MRI or density changes on CT as the clinical endpoint for RBE modeling and found these changes to be dependent on LET. Peeler *et al.* observed that MRI changes depended on increasing LET and dose in patients with pediatric ependymoma. Bertolet *et al.* observed a similar association in meningioma patients treated with pencil beam scanning proton therapy (13, 26). Engeseth *et al.* found that dose and LET were significantly correlated with radiation-associated changes in brain imaging in patients treated for head and neck cancers (27). Underwood *et al.* studied voxel-level CT imaging changes in the lungs of 20 postmastectomy patients receiving proton therapy or photon therapy (28) and found that those receiving proton therapy had a greater increase in both image density and IDC. The authors found that a higher IDC per dose was associated with higher-grade abnormalities, suggesting clinical RBE values above 1.1. There is currently no consensus on the exact relationship between RBE and radiation-induced toxicity (29–31); however, our results indicated a linear correlation between RBE and LET_d_. Several other studies have found correlations between higher RBE and LET with MRI changes, indicating that imaging modalities other than CT could be used to measure RBE (32–34).

In addition, fractionation schemes may also have an important role in determining lung imaging response. In the present study, we found significant imaging changes in PSPT patients compared with IMRT patients, whereas Li *et al.* did not observe significant imaging responses between patients receiving photon therapy and those receiving proton therapy as part of stereotactic body RT until approximately 9 months after completion of RT(35). It is plausible that, given the higher doses per fraction used in SBRT, the RBE is closer to 1. Further studies are needed to compare image-derived RBE in the context of different fractionation schemes and later endpoints.

Our results from validating the measured RBE values indicated good correlation between the different models and demonstrated the feasibility of using non-invasive metrics as a valuable clinical endpoint to calculate RBE. Of the four models, the Mairani model had the worst correlation, possibly because the model includes only LET_d_ in the RBE calculation, whereas the other models include both dose and LET_d_ (24). The measured RBE values were also higher than those derived from the four empirical models. This was likely in part a result of these empirical models being derived from datasets that consisted mostly of survival data for tumor cell lines and in part a consequence of the dose-averaging of LET (9, 23, 25). The model was further validated by fitting two published empirical models. In fitting the McNamara and Wedenberg models, we found the measured RBE to be smaller than values calculated from the fitted model with larger uncertainties. This variability maybe due to the underlying assumption made in deriving each model. For example, in the derivation of the McNamara model, p_0-3_ are free parameters to fit the RBE at 2 Gy, with the assumption that proton α/β varies with LET, that p_0-_ and p_1_ are related to the α term for defining RBE_max_ (where dose approaches 0), and that p_2_ and p_3_ are related to the β term for defining RBE_min_ (high dose limit) (9). However, we do not make these assumptions in modeling the in vivo RBE model. Recent work by Gardner et al. in benchmarking 13 different phenomenological RBE models demonstrated that the underlying assumptions and large uncertainties on the experimental data can cause significant disagreement between models and (36). The results from benchmarking the measured RBE to ones derived from empirical models agrees with the finding from Gardner et al. and highlights the need in vivo specific RBE models.

There are limitations to this work. The dose and image voxels were resampled into “super” voxels to account for DIR uncertainty, which could have affected the observed imaging changes in regions that did not receive radiation. By using these large voxels, we could have captured other biological responses, such as damage to nearby vessels, an abscopal effect or infection that might account for the observed imaging changes. Another limitation of our work was that our dose-response model was fitted to input values obtained by averaging voxel-level values first within patient and then across patients, thus not fully capturing inter-patient variability. Future work should further explore how to integrate patient variability and spatial relationship between voxels into the model. The work presented here was limited to normal tissue; however, this analysis can be extended to examining RBE in tumor tissue. This approach however can be extended to physically tracking voxels; for example, it could be used to create a map showing voxels’ proximity to the tumor surface, which could provide more insight into the varying sensitivity of tissue voxels in biological dose damage relative to the tumor. Another limitation was the large variability in the follow-up scans, which could have affected the magnitude of the radiographic changes observed, as these imaging changes become more apparent 2-4 months after RT (37). Although we found no significant variation within the cohort, future studies should consider how longer durations between the planning CT and follow-up scans. The patients included in this study received RT with standard fractionation (2 Gy per fraction), which is consistent with the data used to fit the empirical RBE models; however, other radiation schemes, such as SBRT, are also used for cancer treatment. Future work should consider how these different factors may affect the measured RBE and how it compares to that determined with different empirical models.

## Conclusion

Using CT-based IDC as the clinical endpoint, we obtained voxel-level variable RBE values for patients with LA-NSCLC who received IMRT or PSPT. Our findings demonstrate the feasibility of using clinical imaging to model proton RBE and warrant additional studies of similar approaches to further elucidate the biological effects of proton therapy.

## Data Availability

The data that support the findings of this study are available from the corresponding author upon request. The data are not publicly available owing to privacy and/or ethical restrictions.

## Acknowledgments

We acknowledge the Editing Services, Research Medical Library at MD Anderson Cancer Center.

---

**Supplementary Figure 1.** A and B, Box and whisker plots of dose (A) and image density change (IDC; B) for each cohort. Boxes indicate the 25^th^–75^th^ percentiles, horizontal lines indicate the medians, plus signs (+) indicate the means, and whiskers indicate the ranges.

**Supplementary Figure 2.** Statistical characterization of individual patient LET in the PSPT cohort.

**Supplementary Figure 3.** Patient level IDC and dose relationship.

**Supplementary Figure 4.** Comparison of RBE value of each model as a function of dose.

