## Supplement for "Clinical evidence of variable proton relative biological effectiveness in locally advanced non–small-cell lung cancer patients"

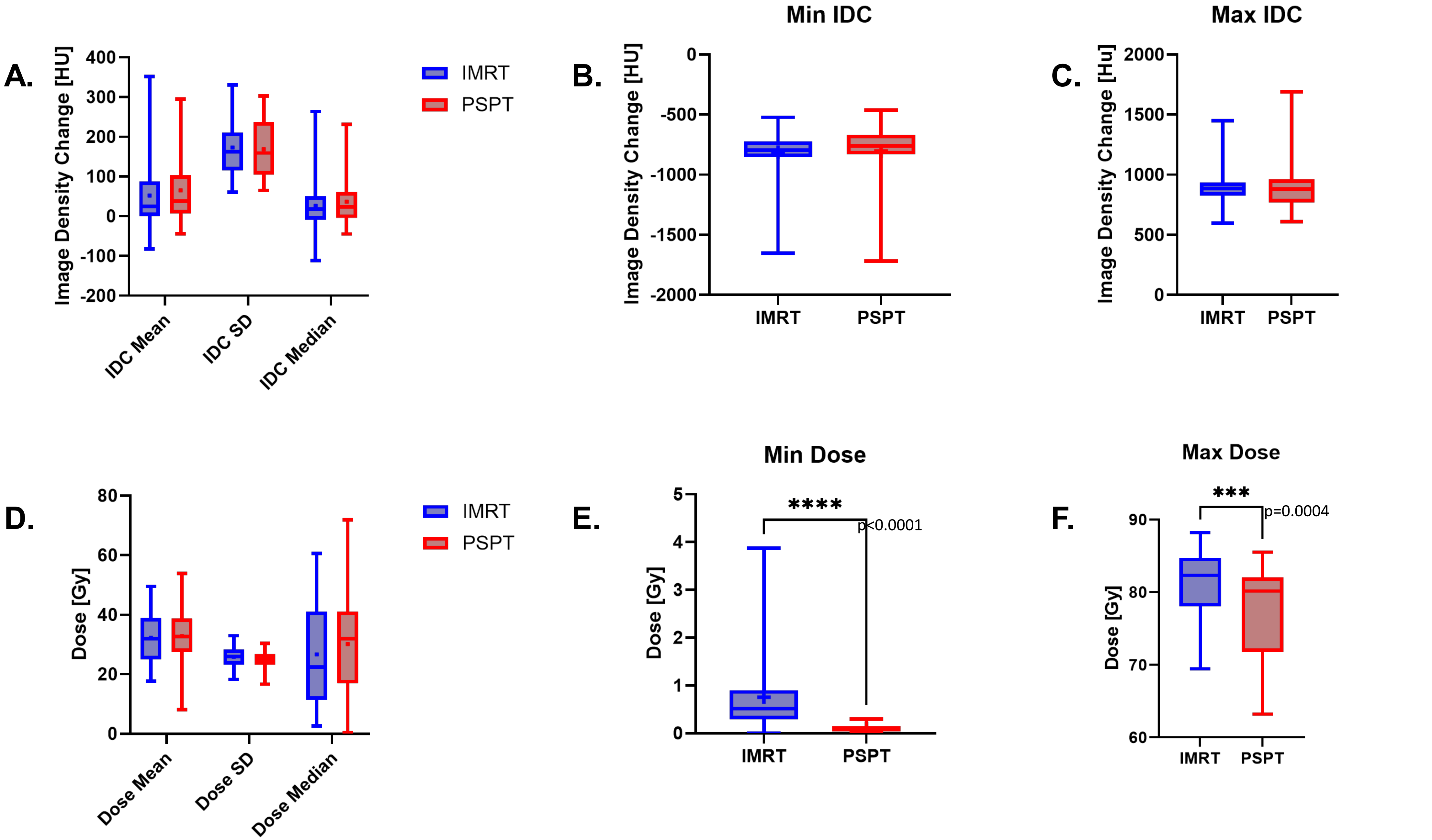


**Supplementary Figure 1.** A and B, Box and whisker plots of dose (A) and image density change (IDC; B) for each cohort. Boxes indicate the 25th–75th percentiles, horizontal lines indicate the medians, plus signs (+) indicate the means, and whiskers indicate the ranges.

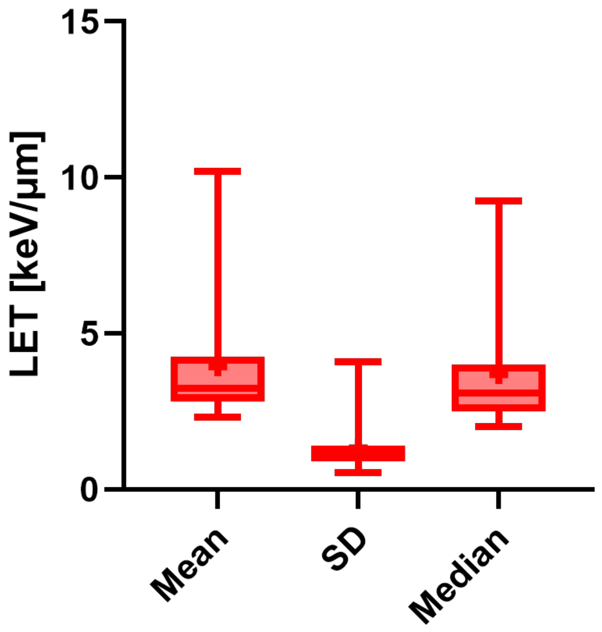


**Supplementary Figure 2.** Statistical characterization of individual patient LET in the PSPT cohort.

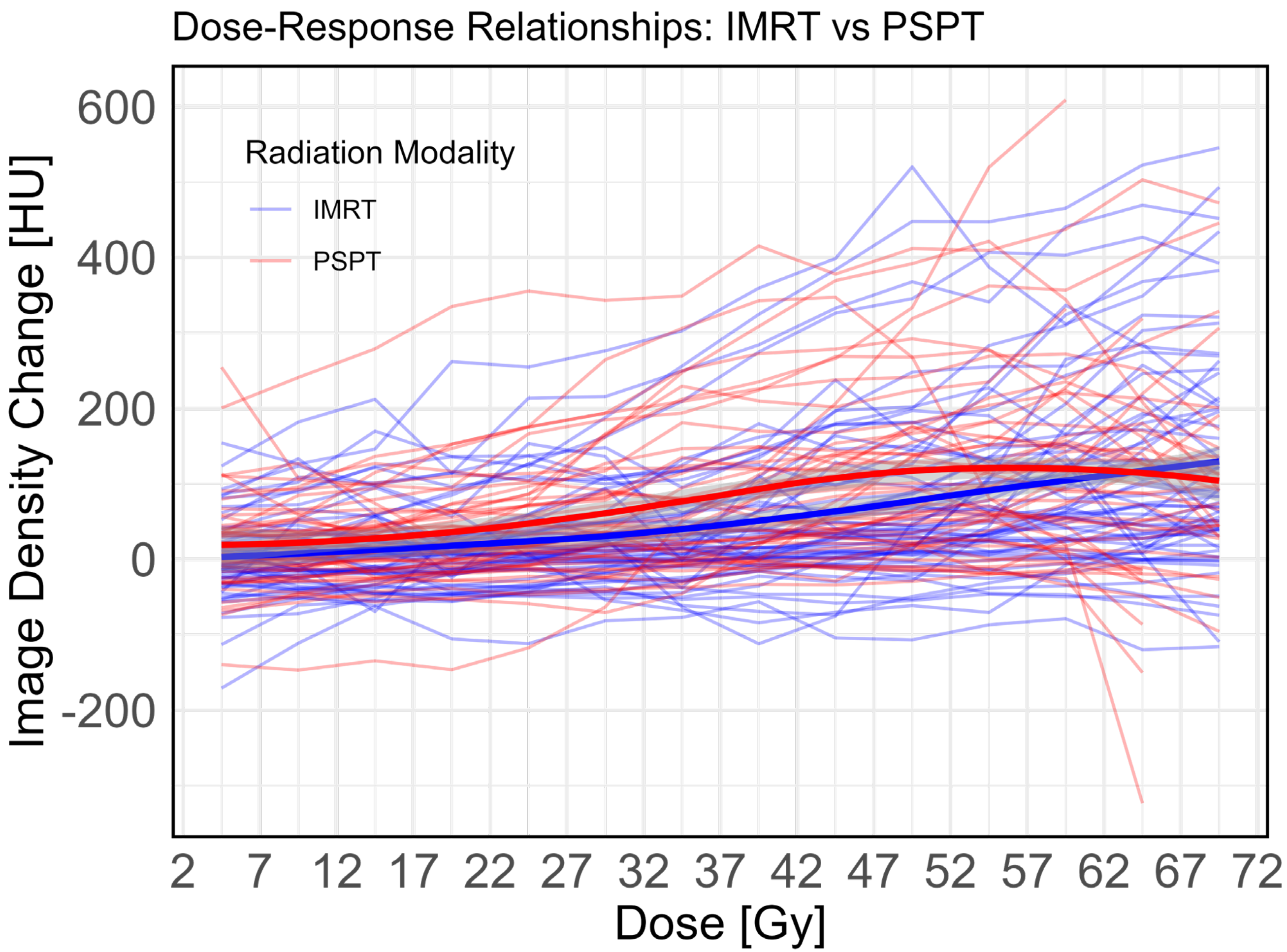


**Supplementary Figure 3.** Patient level IDC and dose relationship. Weighted line represents a smoothed conditional mean.

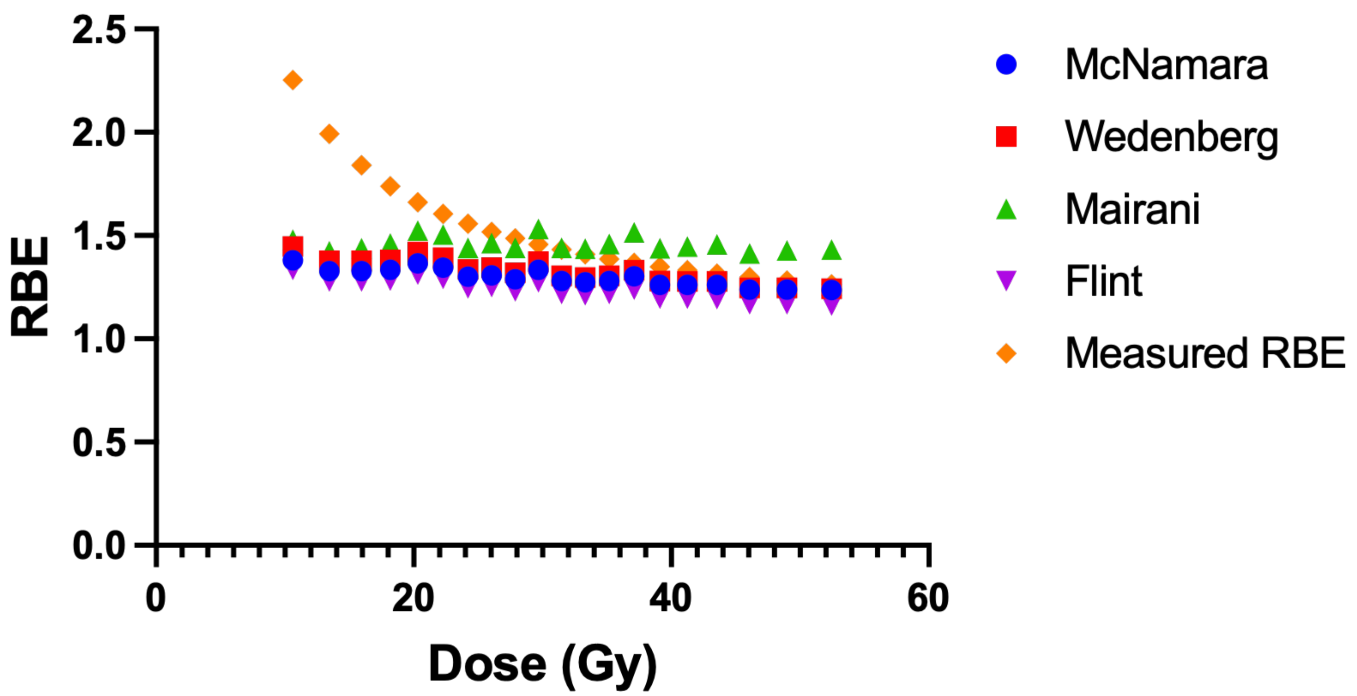


**Supplementary Figure 4.** Comparison of RBE value of each model as a function of dose.
